# Implementation of an interactive mobile application to pilot a rapid assay to detect HIV drug resistance mutations in Kenya

**DOI:** 10.1101/2021.05.06.21256654

**Authors:** Justin D. Vrana, Nuttada Panpradist, Nikki Higa, Daisy Ko, Parker Ruth, Ruth Kanthula, James J. Lai, Yaoyu Yang, Samar R. Sakr, Bhavna Chohan, Michael H. Chung, Lisa M. Frenkel, Barry R. Lutz, Eric Klavins, Ingrid A. Beck

**Affiliations:** Department of Bioengineering, University of Washington, Seattle, Washington, USA; Global Health of Women, Adolescents, and Children (Global WACh), School of Public Health, University of Washington, Seattle, Washington, USA; Center for Global Infectious Disease Research, Seattle Children’s Research Institute, Seattle, Washington, USA; Paul G. Allen Center for Computer Science & Engineering, University of Washington, Seattle, Washington, USA; Department of Electrical and Computer Engineering, University of Washington, Seattle, Washington, USA; Coptic Hope Center for Infectious Diseases, Nairobi, Kenya; Center for Virus Research, Kenya Medical Research Institute, Nairobi, Kenya; Department of Global Health, University of Washington, Seattle, Washington, USA; Department of Medicine, Emory University, Atlanta, Georgia, USA; Departments of Global Health, Medicine, Pediatrics, and Laboratory Medicine, University of Washington, Seattle, Washington, Washington, United States

**Keywords:** HIV drug resistance, rapid assay, implementation, mobile application, laboratory operating system, oligonucleotide ligation assay, low-resource settings

## Abstract

**Introduction:** Usability is an overlooked aspect of implementing lab-based assays, particularly novel assays in low-resource-settings. Esoteric instructions can lead to irreproducible test results and patient harm. To address these issues, we developed a software application based on “Aquarium”, a laboratory-operating system run on a computer tablet that provides step-by-step digital interactive instructions, protocol management, and sample tracking. Aquarium was paired with a near point-of-care HIV drug resistance test, “OLA-Simple”, that detects mutations associated with virologic failure. In this observational study we evaluated the performance of Aquarium in guiding untrained users through the multi-step laboratory protocol with little supervision.

**Methods:** A feasibility study was conducted in a laboratory at Coptic Hope Center in Nairobi, Kenya. To evaluate the training by Aquarium software, twelve volunteers who were unfamiliar with the kit performed the test on blinded samples (2 blood specimens; 5 codons/sample). Steps guided by Aquarium included: CD4+ T-Cell separation, PCR, ligation, detection, and interpretation of test results. Participants filled out a short survey regarding their demographics and experience with the software and kit.

**Results and discussion:** 12/12 laboratory technicians had no prior experience performing CD4+ separation and 7/12 had no experience performing laboratory-based molecular assays. 12/12 isolated CD4+ T cells from whole blood with yields comparable to isolations performed by trained personnel. The OLA-Simple workflow was completed by all, with correct visual and software interpretation of results for 90% (108/120) and 97% (116/120) of codons, respectively. In the surveys, participants favorably assessed the use of software guidance.

**Conclusions:** Aquarium digital instructions enabled first-time users in Kenya to complete the OLA-simple kit workflow with minimal training. Aquarium could increase the accessibility of laboratory assays in low-resource-settings and potentially standardize implementation of clinical laboratory tests.

## Introduction

In resource-rich communities, automation drastically improves the daily operation of clinical laboratories [1–3]. Patient samples can be quickly shipped to centralized laboratories for batch-processing using highly-efficient workflows that generate high-quality results while reducing costs and turnaround time [4, 5]. However, total automation is ill-suited for low-resource settings for many reasons. First, shipping of samples to centralized laboratories can take ≥10 days [6], which undermines the benefits of fast turnaround test results from an automated workflow. Second, in small communities, the demand of a clinical assay may be low, requiring a longer waiting period to receive enough samples to complete a full batch. Finally, automation is often used in conjunction with high-throughput robotic equipment that is cost-prohibitive for small laboratories. In low-resource settings, high-quality and fast laboratory results for complex assays will likely require unorthodox approaches to automation to build on low-cost equipment and be applicable for small batches of samples.

HIV infects nearly 40M people globally [7] and successful management of HIV relies on multiple laboratory tests. Recent advances include point-of-care HIV diagnosis and viral load quantification [8, 9]. Due to the complexity of HIV drug resistance (**HIVDR**) tests used to guide treatment regimens, they are performed in centralized, highly-equipped laboratories [10–13]. In low-resource countries with high HIV prevalence like Kenya, few laboratories have the capacity to test for HIVDR [14]. For laboratories without access to sequencers, an oligonucleotide ligation assay (**OLA**) has been implemented [15] but onboarding OLA required extensive training due to its complexity.

We envision the use of software to automate a simplified version of OLA that uses low-cost equipment. To that end, we developed “OLA-Simple” which uses lyophilized reagents to simplify the workflow and lateral flow tests to provide visual results [16–18]. In addition, we developed a software application based on “Aquarium” [19] that employs human-in-the-loop automation to tightly integrate all the steps in OLA-Simple. Aquarium provides step-by-step interactive digital instructions, protocol management, data collection and sample tracking. In a pilot study at the University of Washington, Aquarium enabled minimally-trained students to accurately perform the OLA-Simple workflow [18]. Here, we demonstrate the use of the Aquarium-enabled HIVDR test in a small laboratory in Nairobi, Kenya.

## Methods

### Laboratory setup at Coptic Hope Center in Nairobi, Kenya

A Seattle team travelled to Nairobi in April 2018 to set-up a testing site at the Coptic Hope Center for Infectious Diseases, which is a large-scale, antiretroviral treatment site [20]. The laboratory’s existing standard thermal cycler, biological safety hood, bench space, and refrigerator were utilised. To onboard the test, the team brought the OLA-Simple kits, minicentrifuge, micropipettes, scanner (CanoScan LiDE 300), and tablets (Fire HD), foot pedal, UPS battery backup and surge protector (APC 1500VA Compact), and server (Intel NuC NUC7i3BNH Mini PC/HTPC) to set up and run Aquarium. Aquarium code is publicly available [21].

### Study design

This feasibility study was designed to evaluate the utility and performance of Aquarium in guiding first-time users to perform the OLA-Simple kits. We recruited 12 laboratory technicians from the Coptic Hospital clinical laboratories to perform OLA-simple following the instructions provided by the Aquarium-based application from April 4 – 13, 2018. Considering the timeframe and resources, we estimated this sample size was adequate to assess the test performance and obtain feedback on the participants’ perceptions related to the OLA-Simple kit and Aquarium digital instructions. This study was approved by the Institutional Ethics Review Committee (IERC) of the Aga Khan University in Kenya and Seattle Children’s Research Institute’s IRB.

### Evaluation of OLA-Simple by participating laboratory technicians

Testing was spread over six days (two techs/day) and consisted of completing a demographic questionnaire and a 30-minute introduction of the kit principle and procedure, followed by processing and testing of two blinded blood samples. CD4+ cells were separated from 0.5 mL uninfected blood and lysed. The cell lysates were then spiked with mixtures of plasmids containing known HIV drug resistance mutations and amplified by PCR, followed by ligation of mutation-specific probes and detection of the ligated products using lateral flow strips. The lateral flow strips were scanned, and the images displayed on the tablets were used by the participants to make visual calls and generate a report using Aquarium. Finally, the participants completed a questionnaire to give feedback on their experience with the kits and software.

### Preparation of OLA-Simple kits

The OLA-Simple kit was prepared and assembled as previously described [22]. The EasySep isolation kit (STEMCELL Technologies, Vancouver, CA) to negatively select CD4+ T-Cells was adapted to small blood volume processing, aliquoted and packaged in foil pouches. Reagents for PCR, ligation for detection of five HIV major NNRTI/NRTI resistance codons (K65R, K103N, Y181C, M184, and G190A) and lateral flow strips to detect ligation products were packaged in foil pouches with desiccant. Each kit component was labeled with a unique identifier, matching the images illustrated in Aquarium instructions.

### Post-analysis of samples in Seattle

The DNA yield in lysed cells obtained by Kenyan participants was assessed in Seattle by qPCR of human beta globin [23]. The lateral flow images and Aquarium reports generated by each participant were used to assess assay performance and interpretation of visual results. Test accuracy was determined by comparison to the expected genotype at each codon analysed. Scanned images of lateral flow strips were re-analysed using an in-house Python script [22] to determine if automated analyses improved test accuracy. 95% confidence intervals (CI) are reported for all proportions.

## Results and discussion

This work presents the first use of human-in-the-loop automation to enable a resource-limited laboratory to successfully operate an HIVDR test with minimal training. We developed a custom mobile application based on Aquarium operating system that describes procedures and workflows for our OLA-Simple kits. Here we describe the performance of the OLA-Simple kits in the hands of first-time users in a small laboratory in Kenya and their feedback on their experience using the Aquarium digital guidance.

### Participant characteristics

The 12 participants recruited were 83% male, median age 30 years old (range 26-42), had a median of 6 (range 3-10) years of experience as a laboratory technician, and all were conversant in English. Their education level ranged from secondary school with a certificate in Medical Laboratory Technology to a Master’s degree in a Laboratory Science field, which is representative of most clinical laboratories in Kenya. They reported varied levels of experience working with HIV or molecular techniques (**Table 1**).

**Table 1.**
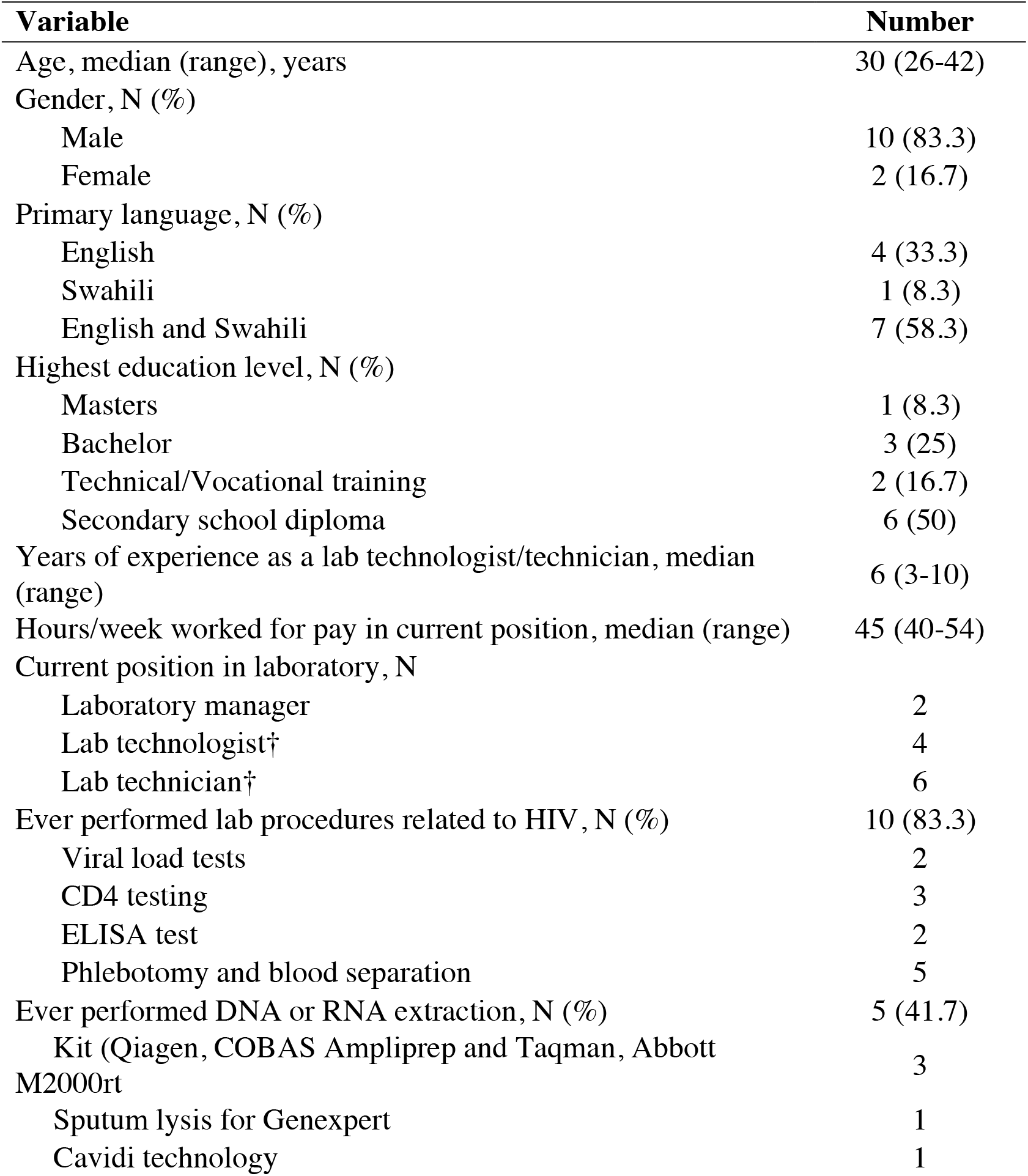
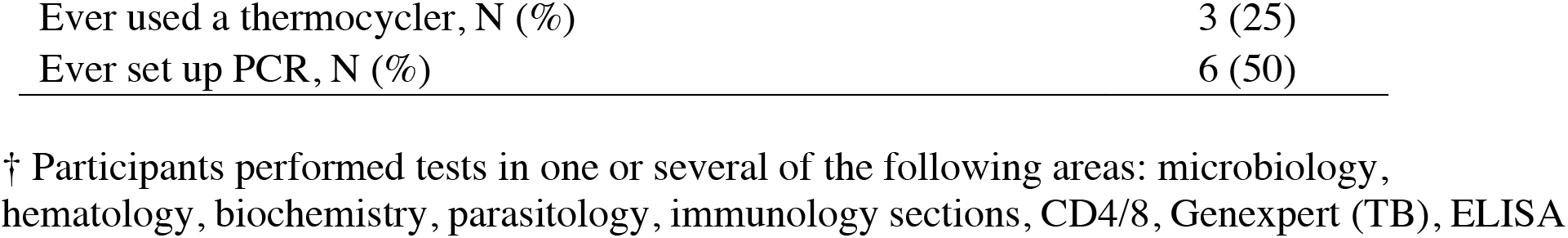
Demographics of participants (N=12)

### Laboratory setup for Aquarium-assisted OLA-Simple training

The installation cost of OLA-Simple is low compared to automated sequencing platforms (> US$ 100,000). The workflow uses equipment that exists in most laboratories or is relatively inexpensive to acquire. Setting up the assay and software at the Coptic Hope Center laboratory required ∼US$1,000 of additional equipment (scanner, microcentrifuge, vortexer, computer tablets, a server, and an uninterruptible power supply). For a laboratory without access to a thermal cycler, a battery-powered portable unit (∼$500) can be used as we have previously shown [24].

The laboratory was set up with a Wi-Fi network to run Aquarium and coordinate assay steps across a pre-PCR and post-PCR room (**Figure 1A**). Participants worked in pairs following the implementation workflow (**Figure 1B**): sample preparation and PCR set-up were carried out in the pre-PCR room, while PCR, ligation and detection were conducted in the post-PCR room to minimise potential for amplicon-carryover contamination. Each participant processed two uninfected blood specimens and performed mutation testing on two contrived specimens with known HIV mutations following the interactive digital instructions provided by Aquarium.

**Figure 1.**
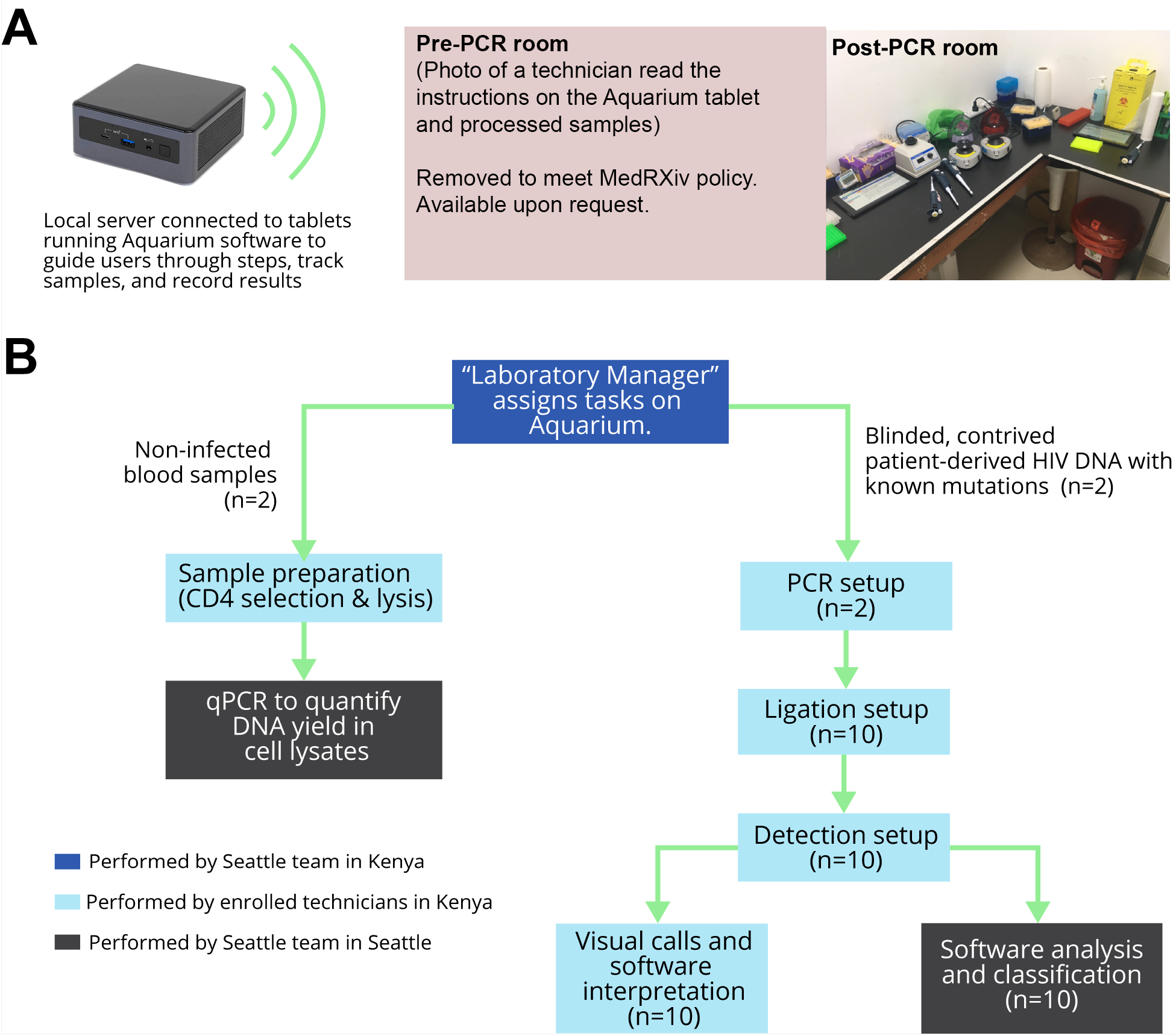
OLA-Simple laboratory setup and workflow. (**A**) Laboratory setup: Tablets were connected to a local server to run Aquarium software. The pre-PCR room had a refrigerator/freezer and a BSC where CD4+ separation and PCR reaction were set-up. Technicians controlled the Aquarium-based software on the tablet with a foot pedal (not shown in the picture) while performing CD4+ preparation. The post-PCR room had two designated bench areas to set up ligation and detection separately. This room also contained a thermal cycler and a scanner. (**B**) Tasks assigned by Aquarium for Kenyan technicians to perform, and assessment of their performance using Aquarium-assisted OLA-Simple. The sample preparation module was separated from the amplification, ligation, detection, and interpretation module. Documentation, associated protocol code, and examples of complete runs are publicly available (https://github.com/OLA-Simple/Papers-Vrana-Panpradist-et-al-2021).

On average, it took seven hours for a pair of participants to complete the tasks in the workflow from introductory session to completion of surveys. This turnaround time also included staggering the work of two participants due to space and instrument constraints and would likely decrease to 4.5-5 hours once users became familiar with the software and the kits.

Reported turnaround time for centralised HIVDR testing in resource-limited settings is 18 days [25] from sample collection, shipping to the laboratory, processing of samples, and transmission of results to the clinic. In this study, OLA-Simple was performed in a laboratory located within the hospital grounds and thus could deliver test results within one day. In addition, each kit tests two samples, a number suitable for the volume of weekly patient samples submitted for HIVDR testing in small laboratories in Kenya, which eliminates the need to batch specimens for testing.

### Performance of participants using OLA-Simple

All 12 participants successfully isolated CD4+ cells from whole blood collected from four donors with yields within the expected range (mean±SD: 686,450±216,500 CD4+ cells/mL, as determined by qPCR) and completed genotyping of two samples using the OLA-Simple kit reagents and protocols as instructed by the Aquarium application. The two blinded DNA samples tested by all participants included wild-type genotype only or mixtures of mutant and wild-type genotypes at each of five HIV reverse transcriptase codons tested by OLA-Simple: K65R, K103N, Y181C, M184V and G190A (total of 10 codons/participant) (**Figure 2A**). Participants correctly genotyped 70/72 (97.2%, 95% CI: 90-100%) mutant codons and 38/48 (79.2%, 95% CI: 65-90%) wild-type codons. The only two false negatives were due to Participant #6 erroneously testing Sample 1 twice and omitting testing of Sample 2. Of 10 false positive results, 2 were due to testing Sample 1 in place of Sample 2 by Participant #6, 1 appeared to be contamination (light mutant signal at codon G190A likely from reusing a pipette tip contaminated with mutant ligation product) and 7 were due to light mutant background at codons K103N (n=6) and K65R (n=1). Analysis of the scanned images using our in-house image analysis software improved test accuracy to 97% (116/120, 95% CI: 92-99%) with 97% (70/72, 95% CI: 90-99%) mutant codons and 96% (46/48 95% CI: 86-99%) wild-type codons correctly genotyped (**Figure 2B**).

**Figure 2.**
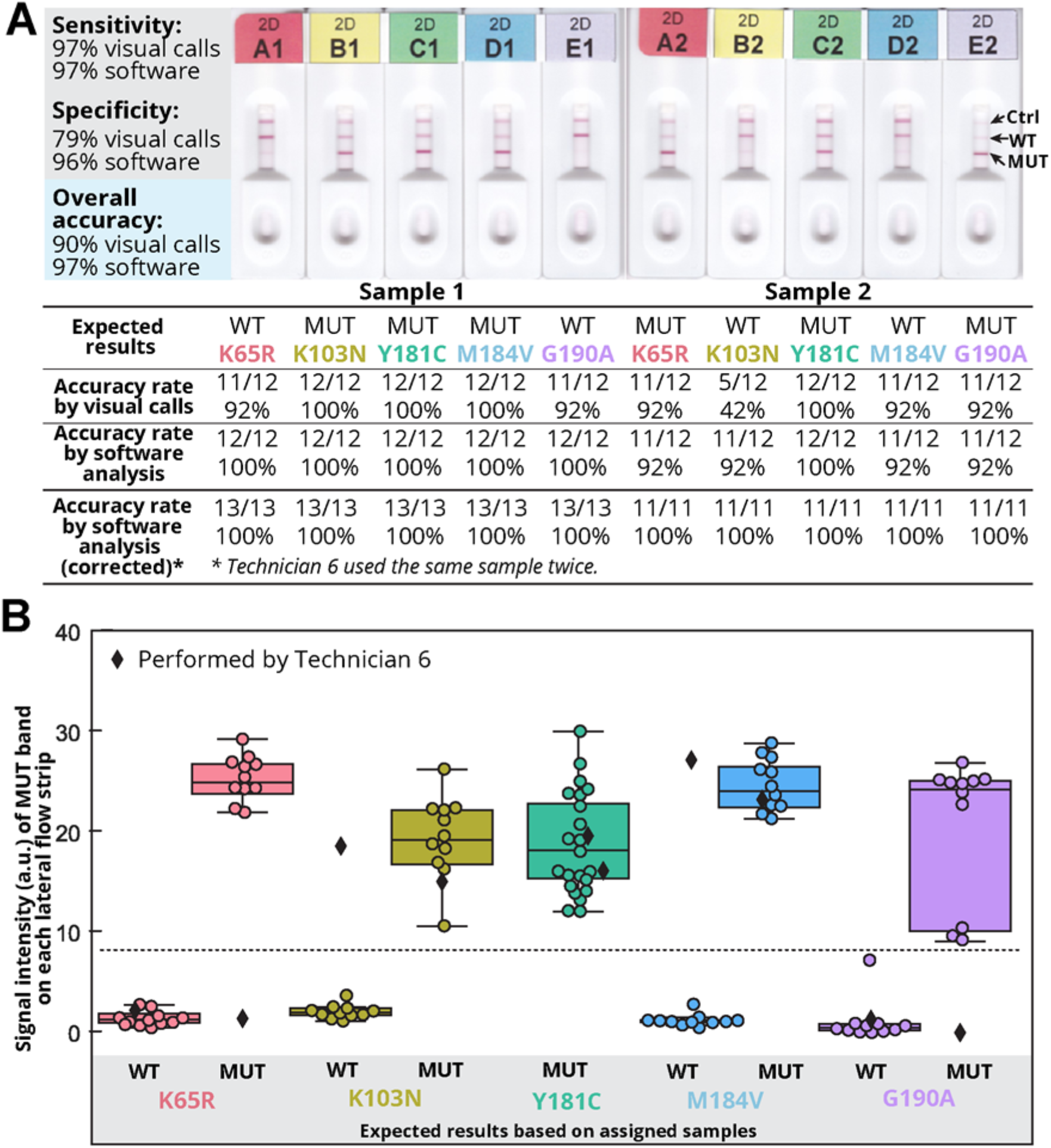
OLA-Simple HIVDR results and interpretations. (**A**) Examples of scanned images of test strips for Samples 1 and 2 and test accuracy across each codon based on visual calls made by each participant and post-processing by image analysis software. Sample1 and Sample 2 have different mutation profiles. (**B**) Mutant (MUT) signal intensity of each lateral flow strip. Middle lines on the box plot indicate medians. Top and bottom lines on box plot indicate interquartile ranges. Dashed lines indicate the detection threshold for MUT signal. Both Sample 1 and Sample 2 have mutant genotype at codon Y181C. Diamonds correspond to signal from the strips of a Sample 1 that was erroneously tested in place of Sample 2.

The image analysis software improved accuracy over visual interpretation of results by establishing a signal threshold above the mutant background signal and thus eliminating false positives. The sample mix-up described above reduced the overall accuracy; this could be improved with changes in the kit labeling system to prevent this type of errors. Correcting for the sample that was added twice, the performance of the assay chemistry combined with software analysis, yielded 100% accuracy (120/120, 95% CI: 97-100%) with correct results for 100% (72/72, 95% CI: 95-100%) mutant codons and 100% (48/48 95% CI: 93-100%) wild-type codons.

### Feedback on software and kit features

Overall, participants scored the use of software as helpful in learning to perform the assay. Qualitatively, participants enjoyed the clear instructions and interpretability of the results using the tablets. They strongly agreed that they understood the meaning of the bands on the strips, and that the Aquarium instructions were easy to follow. However, several participants felt the procedure was lengthy and involved too many steps (**Supplementary Tables 1 and 2** summarise the survey responses). In response, we have subsequently developed new chemistries to reduce assay time to 3.5 hours by replacing the 1.5-hour blood DNA preparation by a 30-minute plasma RNA extraction and the 2-hour PCR by 1-hour RT-PCR.

Our study shows it is feasible to use Aquarium to train local laboratory personnel with basic experience in lab work and onboard an HIVDR test, but this pilot study was limited to the use of contrived specimens to establish analytical performance. Due to limited resources and allocated study time, each participating laboratory technician was able to perform the OLA-Simple kit once. Thus, we were not able to assess the ability of each technician to maintain or improve their skills. A larger demonstration and evaluation study that includes processing HIV-infected clinical specimens on multiple OLA-Simple runs is ongoing in Kenya to assess the robustness of our HIVDR test.

The Aquarium software has useful features for HIVDR testing such as automatic collection of operator interactions in Aquarium’s virtual laboratory notebook that can be useful for troubleshooting. Each kit item is labeled with a unique identifier that Aquarium instructions use in conjunction with corresponding pictures to avoid ambiguity and reduce the extent of in-person training needed. Uniquely labeled items also allow tracking of stock consumption in real-time which can be useful and timesaving for laboratory management. Importantly, Aquarium could link test results to treatment algorithms to advise clinicians, and algorithms could be changed as clinical recommendations or policies change.

## Conclusion

Aquarium-based software enabled deployment of the OLA-Simple with minimal training by lowering technical skills required to perform such test. Local laboratory technicians operated the OLA-Simple for the first-time with good recovery in sample preparation and high accuracy of HIVDR detection. Human-in-the-loop automation could facilitate daily operations of laboratory-based assays and increase the accuracy and assay performance in small laboratories.

## Data Availability

All data were available in the main text and supplementary information.

## Authors’ contributions

NP, RK, and YY conceived the vision for Aquarium for OLA-Simple. NP, NH, DK, RK, IB, and JL developed and prepared the OLA-Simple kits. JDV, YY, and EK developed the software application for this work. JDV and IB travelled to Kenya to conduct this study. IB, NP, PR, and JDV analysed the data. BRL, LMF, EK oversaw the overall study on the Seattle side. SRS, BC and MHC oversaw the study design on the Kenya side and helped enrolled local technicians in Kenya. All authors contributed to writing of this manuscript and approved the final version for submission.

## Acknowledgments

We are grateful to the Coptic Hospital and the Coptic Hope Center for their support and to the technologists from these institutions who participated in this study. We thank Syamal Raychauduri, Dindo Reyes, Kim Polizzi, and Jessica Price at InBios International, Inc. for producing the lateral flow strips used in the validation study; and Aarthy Vallur and Kathryn Hjerrild of InBios International, Inc. for providing feedback on OLA-Simple prototypes.

