## Supplementary Tables for "Implementation of an interactive mobile application to pilot a rapid assay to detect HIV drug resistance mutations in Kenya"

**Supplementary Tables. Survey responses from participants regarding Aquarium and the OLA-Simple kit.**

Participants were asked to fill out a short survey immediately following the procedure, which included evaluating four statements on a 5-point Likert scale (strongly agree to disagree) (Supplementary **Table 1**) and three open-ended qualitative questions (Supplementary **Table 2**). For ordinal categorical values, modes are displayed for each statement. For open-ended questions, we present a summary of the responses describing the different points raised by all the participants.

**Table 1. Responses to statements**

| **Questionnaire statement** | **Mode** |
|---|---|
| **Specimen preparation**: I was able to perform these steps in less time than my usual DNA extraction | 4 |
| **PCR and Ligation***: Using dried reagent is easier than setting up a traditional PCR reaction | 4 |
| **Detection**: I understood the meaning of the bands in the strip | 5 |
| **Kit instructions**: Instructions were easy to follow | 5 |

* Participants 11 and 12 answered NA to the PCR question, likely due to not having performed PCR prior to this training

Rating scale: 1=strongly disagree, 2=disagree, 3=neutral, 4=agree, and 5=strongly agree.

**Table 2**. **Summary of responses to open-ended questions**

| What did you like best about using this kit? | Which instruction(s) were not easy to follow? Please explain why. | What advice do you have for us to make kit and instructions easier to use? |
|---|---|---|
| • The instructions were easy to follow<br>• The protocol was easy to understand.<br>• The examples in Aquarium were well labeled and made it easy to know where to add each reagent or sample<br>• The kit is user-friendly and straightforward<br>• The kit is easy to use compared to the plate-based OLA method<br>• There was only one PCR step instead of the usual two for nested PCR<br>• Detection was simplified by Aquarium<br>• Results are generated fast and are also portable<br>• It is easy to read the bands and interpret the results: it clearly shows the control, the wild-type and the mutant<br>• A single sample can be easily processed without the need for batching | • The instructions and procedure were clear and easy to follow.<br>• The instructions were Ok when following a step, but it was difficult to memorize the steps without a written protocol to follow<br>• Some instructions did not include the specific vial or reagent in the heading, but is was in the pictorial<br>• Sample preparation:<br>- the multiple tubes and SOP instructions were not alphabetically arranged<br>- includes many tubes, color coding would help to identify which to use first and avoid mix-ups<br>- too many steps make it confusing<br>- the process is long with many reagents and waiting time<br>• During the ligation/detection step had challenges remembering to add reagent to the next strip<br>• At times the tablet was not working, and we did not have a backup plan such as a written protocol with instructions. | • Enlarge the font under every heading.<br>• Include the reagent vial ID on the headings for better clarity<br>• The tube labels should match the SOP instruction labels.<br>• Harmonize the sequence of adding reagents and closing of tubes to reduce multitasking and possible confusion.<br>• Create system for how to keep track of addition of reagents to tubes<br>• State what to expect at the end of each step, e.g. maybe a clean supernatant /deposit so one can confidently move to the next step<br>• Look for alternatives to replace the current sample preparation method<br>• Make the incubation time shorter<br>• Improve turn-around time for clients.<br>• Have print outs of the protocols as a back-up plan.<br>• Make the kit commercially available with all the steps for laboratory technologists to follow. |
